# Moderators of Treatment Outcomes for LGBTQ+ Military Veterans in the PRIDE in All Who Served Health Promotion Group

**DOI:** 10.1101/2023.02.15.23285954

**Authors:** Michelle M. Hilgeman, Robert J. Cramer, Andréa R. Kaniuka, Ryan Robertson, Teddy Bishop, Sarah M. Wilson, Heather A. Sperry, Tiffany M. Lange

**Affiliations:** Research & Development Service, Tuscaloosa VA Medical Center, Tuscaloosa, AL 35404; Department of Psychology, The University of Alabama, Tuscaloosa, AL 35487; Department of Public Health Sciences, University of North Carolina at Charlotte, Charlotte, NC 28233; Research & Development Service, Tuscaloosa VA Medical Center, Tuscaloosa AL; VA Health Services Research & Development Center of Innovation to Accelerate Discovery and Practice Transformation (HSR&D ADAPT COIN), Durham VA Healthcare System, Durham, NC; Department of Psychiatry and Behavioral Sciences, Duke University School of Medicine, Durham, NC; LGBTQ+ Health Program, Veteran Health Indiana; Community Behavioral Health, Philadelphia, PA

**Author notes:** Corresponding Author: Michelle M. Hilgeman, PhD, Research & Development (151), Tuscaloosa VA Medical Center, 3701 Loop Road E., Tuscaloosa, AL 35404. Declarations of Interest: None. Authorship Statements: Hilgeman: Conceptualization, Methodology, Formal Analyses, Writing Original Draft & Reviewing Editing, Supervision, Funding Acquisition Cramer: Conceptualization, Methodology, Formal Analyses, Writing Original Draft & Reviewing Editing, Visualization, Funding Acquisition Kaniuka: Writing Original Draft Robertson: Writing Original Draft Bishop: Project Administration, Data Curation, Writing Original Draft Wilson: Writing Reviewing & Editing Sperry: Writing Reviewing & Editing Lange: Conceptualization, Resources, Funding Acquisition, Writing Reviewing & Editing. Disclaimer: The views expressed in this article are those of the authors and do not necessarily reflect the position or policy of the Department of Veterans Affairs or the United States government.

**Keywords:** minority stress, LGBTQ+, sexual and gender minority, coping self-efficacy, suicide risk, depression, anxiety, military veteran

## Abstract

**Background:** Veterans who identify as lesbian, gay, bisexual, transgender, queer, questioning, and related identities (LGBTQ+) have faced discrimination that puts them at increased risk for depression, anxiety, and suicide. Upstream interventions like the PRIDE in All Who Served program can improve internalized prejudice, suicide attempt likelihood, symptoms of depression, and symptoms of anxiety by addressing minority stress, facilitating social connection, and promoting engagement with the healthcare system. Yet, little is known about who benefits most from these types of services.

**Methods:** Sixty-six US military veterans (Mean age = 47.06, SD = 13.74) provided outcome surveys before and after a 10-week health promotion group for LGBTQ+ individuals at one of 10 Veterans Health Administration (VA) Medical Centers. Coping self-efficacy and key demographic factors were examined as moderators of treatment outcomes.

**Results:** Coping self-efficacy moderated effects across treatment outcomes with those lower in coping self-efficacy beliefs reporting the greatest benefit of the intervention. Reduction in anxiety symptoms was moderated only by problem-solving coping self-efficacy, while suicide attempt likelihood was moderated only by social support. Reduction of internalized prejudice and depression symptoms were moderated by both problem-solving and social support coping self-efficacy, while thought-stopping (a frequent target of traditional cognitive therapies) only moderated internalized prejudice, but not clinical symptom indicators. Most demographic factors (e.g., age, race, gender) did not impact treatment outcomes; however, sexual orientation was significant such that those who identified as bisexual, queer, or something else (e.g., pansexual) had greater reductions in internalized prejudice than their single gender-attracted peers.

**Conclusion:** Individual differences like coping self-efficacy and sexual orientation are rarely considered in clinical care settings when shaping policy or implementing tailored programs. Understanding implications for who is most likely to improve could inform program refinement and implementation of affirming interventions for minoritized people.

---

Who improves from an intervention is a critical question for clinical program evaluation and intervention refinement in healthcare settings (1). This paper examines potential moderators of treatment outcomes for the PRIDE in All Who Served health promotion group for military veterans who identify as lesbian, gay, bisexual, transgender, queer, questioning and related identities (LGBTQ+) in the U.S. Department of Veterans Affairs Health system (VHA; 2, 3). Examining differences in response to this new and spreading program has potential implications for implementation at both the local clinician level and for leadership tasked with meeting needs of an increasingly visible LGBTQ+ veteran population choosing the VHA as their primary place to access health care (4).

## Health Disparities Observed in LGBTQ+ Military Veterans

LGBTQ+ veterans are a marginalized group within the VHA (5). Available estimates suggest there are more than one million veterans who identify as lesbian, gay, or bisexual and approximately 130,000 veterans who identify as transgender or gender diverse (5). Yet, reports from active military service members point to even higher rates, with 6.1% identifying as LGBTQ+ in 2015 (6).

LGBTQ+ veterans face systemic discrimination and other unique barriers to healthcare that impact their access to care (e.g., 7). Inequities in mental health outcomes include higher rates of suicidal ideation and attempts, post-traumatic stress disorder, and depression compared to the general veteran population (8, 9). Worse mental health outcomes are due, in large part, to minority stress (e.g., 10, 11) which includes: (a) experiences of rejection, discrimination, harassment, and victimization, and (b) internalized processes such as internalized stigma, homophobia, biphobia, and/or transphobia. For LGBTQ+ veterans, minority stress stems from both within and outside of the military setting (e.g., 7, 12, 13). These specific minority stressors include the legacy of the Don’t Ask, Don’t Tell policy (14) which was repealed just over a decade ago and banned sexual minorities from serving openly in the military (15), as well as challenges faced by transgender service members and veterans with military bans being lifted and reinstated several times between 2016 and 2021 (16, 17). These policies, while currently rescinded, contribute to a military culture that leads to concealment of LGBTQ+ identity (18), an established contributor to negative mental health outcomes among LGBTQ+ people (e.g., depression, anxiety; 19, 20).

## Health-Promotion Interventions for LGBTQ+ Veterans

Tailored programs that bolster resilience and address the impact of minority stress on LGBTQ+ veterans are emerging in mental health clinics and wellness-focused programs in healthcare settings (e.g., 9). The PRIDE in All Who Served program is an affirmative care intervention developed for delivery outside of mental health settings to promote health for LGBTQ+ veterans at the VA (2). Ten weeks of content address LGBTQ+-related identity resilience and stress (e.g., continuums of identity, identity development, coming out/emergence), enhance health literacy and engagement with services (e.g., sexual health, affirmative care, whole health), increase social connection (e.g., health and safety in relationships, LGBTQ+ community resources), and process minority stress associated with military service experience (e.g., military culture, VA culture). Each session is guided by participant handouts and a manual for group facilitators that includes information on establishing an affirmative environment in the session and more broadly at the facility. Mental health-related diagnoses and related distress are not required for Veterans to access the PRIDE in All Who Served group. Yet, initial pre-post evaluations show reduction in veteran symptoms of distress (e.g., suicidal ideation) and improvement in identity-related resilience like self-acceptance (2, 3). Implementation support is also provided, resulting in positive impacts on facility-level Healthcare Equality Index scores and rapid spread beyond the development site in just five years (3, 21).

## Possible Moderators of LGBTQ+ Intervention Outcomes

The clinical utility of identifying non-diagnostic individual differences that may impact treatment response has received considerable attention (e.g., 22, 23, 24). However, very few studies have examined treatment moderators for interventions created for LGBTQ+ persons. Connecting to the LGBTQ+ community, seeking social support, and coping strategies like cognitive reframing and emotion regulation can buffer the linkage between minority stress and negative mental health (25, 26, 27). The mixed results of prior studies and gaps in empirical literature characterize the current vacuum in which clinicians and healthcare leaders are making treatment delivery decisions for LGBTQ+ individuals in their clinics.

Some demographic variables such as age, gender, and race/ethnicity are widely studied moderators of treatment in mental health outcomes research (28, 29). For example, some review papers have indicated that culturally adapted interventions are more effective when provided to homogenous groups of individuals rather than combining those with mixed cultural, racial, or ethnic backgrounds (e.g., 30, 31, 32), while other individual factors like age and gender have been less consistent. For example, in two studies of PTSD treatment, one found no impact of age while another found an effect of gender with women^1^ reporting greater improvement compared to men (33, 34).

### Coping Self-Efficacy

Coping self-efficacy (CSE) – defined as the beliefs about one’s ability to use coping strategies in the face of aversive experiences – has been linked to mental health and identity-related outcomes in both general population and LGBTQ+ samples (35). Collective evidence suggests CSE and related strategies may be associated with better mental health for LGBTQ+ populations. Coping and resilience buffer the negative impacts of stigma for LGBTQ+ people and are logical mechanisms of focus for intervention (e.g., 36, 37, 38). CSE has also been identified as a mechanism of therapeutic change following cognitive behavioral group therapy for social anxiety disorder (CBGT; 39). After 12-months of CBGT treatment, participants reported greater cognitive reappraisal self-efficacy (i.e., akin to stopping negative thoughts), which was associated with lower levels of social anxiety symptoms. Among LGBTQ+ adults seeking care at an urban Federally Qualified Health Center, lower CSE has been associated with identity concealment and uncertainty about identity, including a decreased belief in the ability to engage in problem-focused coping, thought stopping, or asking for social support (40). Similarly, expectations of interpersonal rejection were related to lower levels of each CSE subscale while higher internalized homonegativity was associated with lower beliefs about using problem-focused coping or ability to receive social support from others in a community-based SM people (41).

A similar pattern of association between domains of coping-self efficacy and improved mental health has been observed in military active-duty samples. Lower levels of thought stopping, problem-focused coping, and asking for social support were linked with greater stress, anxiety, depression, and PTSD symptoms, as well as poorer overall physical health (42). The same study noted greater thought stopping beliefs were associated with less prior (e.g., lifetime suicidal behavior) and future suicidal behavior (e.g., future likelihood of a suicide attempt). Coping self-efficacy beliefs may be of particular importance for veterans due to prior combat-related stressors, such as prolonged threats of danger, isolation from support systems at home, and inability to form community with other LGBTQ+ veterans due to bans on disclosing identity in the military (43, 44). Additionally, access to social support and CSE have been described as protective factors in decreasing PTSD symptoms and overall psychological distress in general veteran population samples (45, 46, 47). Altogether, coping self-efficacy may be a valuable individual difference worth examining within the context of group-based interventions – particularly for LGBTQ+ individuals in clinical settings.

### The Present Study

Very few LGBTQ+-affirming programs for military veterans exist, particularly with evaluation information that includes potential mechanisms (e.g., identity-related resilience; 48). Yet replicability of high-quality programs and data-driven decision-making are core features of a learning healthcare system like VHA (49). PRIDE in All Who Served (2) is an evidence-based promising practice that is increasingly available for LGBTQ+ veterans within the VHA (3, 21). This paper examines clinical program evaluation data from the PRIDE in All Who Served program to identify potential moderators of program outcomes including internalized stigma and related mental health symptoms (i.e., depression, anxiety, and suicidal ideation). Evidence of program outcome moderators may offer opportunities to further improve health promotion programs for LGBTQ+ veterans and identify those who may benefit most.

Therefore, we explored the following questions using clinical program evaluation data:

1. We assessed whether coping self-efficacy would moderate PRIDE in All Who Served outcomes. We hypothesized that those with lower levels of CSE would have greater reductions in internalized prejudice and mental health symptoms over program participation.
2. We assessed whether demographic factors (i.e., race/ethnicity, age, sexual orientation, and gender) would moderate PRIDE in All Who Served outcomes. Given the lack of prior literature on this topic, these analyses were exploratory with no set hypotheses.

## Method

### Participants

Table 1 contains sample demographic and individual difference descriptive statistics. Participants attended PRIDE in All Who Served groups in 2018-2019. Race was primarily White or Black. Sexual orientation, gender identity, and military service branch varied considerably within the sample. The group was primarily of non-Hispanic ethnicity, with an average middle-adult age. Pre-program depressive symptom mean scores were in the moderate range, and post-program scores were in the mild range (50). Both pre- and post-program anxiety symptom average scores were in the mild range (51). Pre-program suicide attempt likelihood average scores fell between “no chance at all” and “rather unlikely,” whereas post-program scores reflected a response of “no chance at all” (52). Both pre- and post-program internalized prejudice mean scores reflected an average response of “strongly disagree” (53). Though typical ranges for coping self efficacy are not established; problem-solving and thought stopping coping self-efficacy average scores were approximately equal to those observed in active-duty treatment-seeking samples, whereas the current sample of LGBTQ+ veterans’ perceptions of their ability to obtain social support was lower compared to published active-duty treatment-seeking samples (42).

**Table 1.**
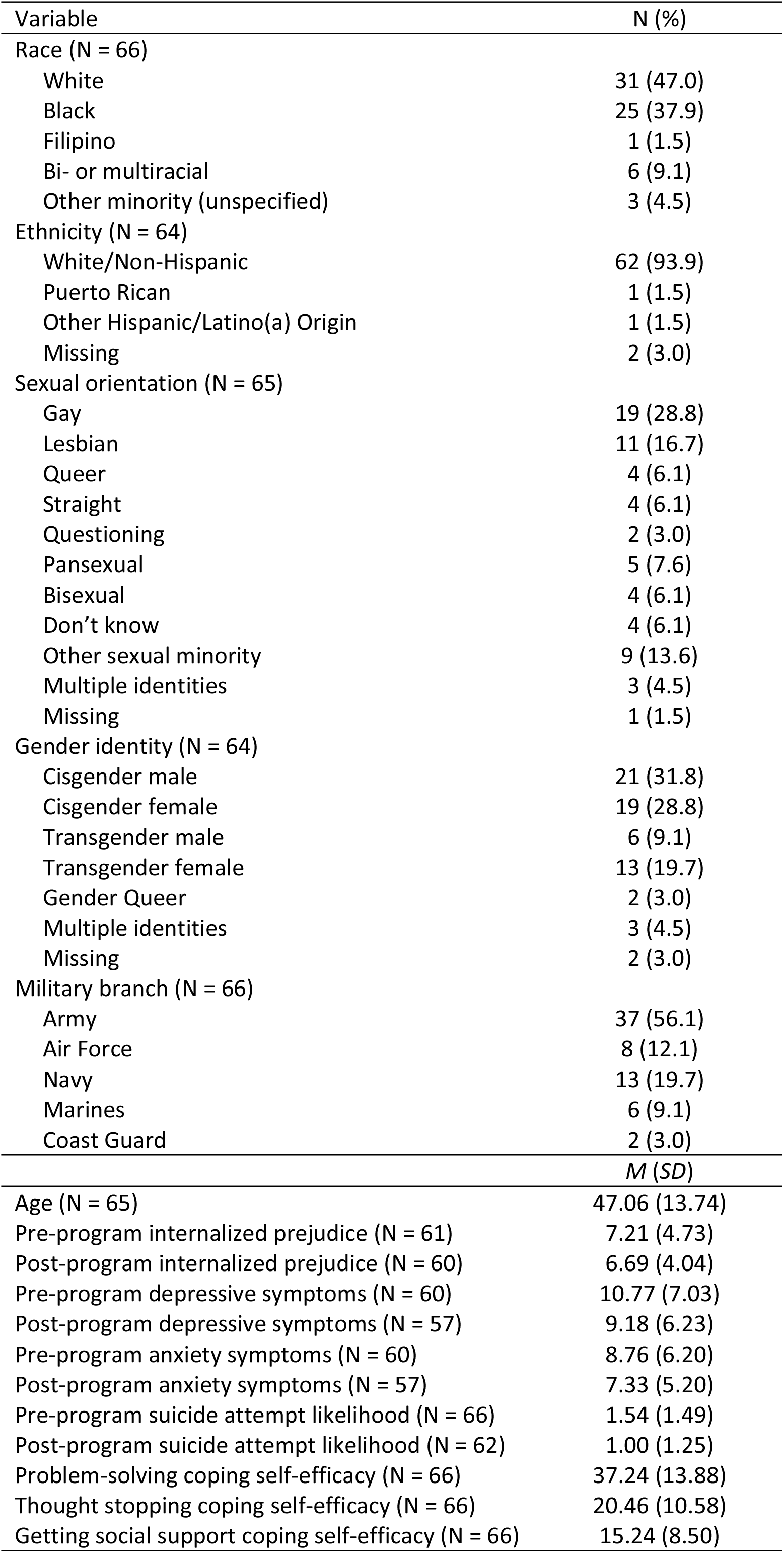
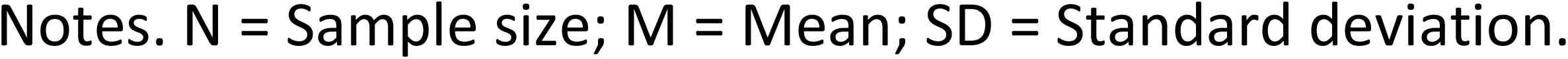
Veteran Demographic and Program Outcome Descriptive Information.

| Variable | N (%) |
| --- | --- |
| Race (N = 66) |  |
| White | 31 (47.0) |
| Black | 25 (37.9) |
| Filipino | 1 (1.5) |
| Bi- or multiracial | 6 (9.1) |
| Other minority (unspecified) | 3 (4.5) |
| Ethnicity (N = 64) |  |
| White/Non-Hispanic | 62 (93.9) |
| Puerto Rican | 1 (1.5) |
| Other Hispanic/Latino(a) Origin | 1 (1.5) |
| Missing | 2 (3.0) |
| Sexual orientation (N = 65) |  |
| Gay | 19 (28.8) |
| Lesbian | 11 (16.7) |
| Queer | 4 (6.1) |
| Straight | 4 (6.1) |
| Questioning | 2 (3.0) |
| Pansexual | 5 (7.6) |
| Bisexual | 4 (6.1) |
| Don't know | 4 (6.1) |
| Other sexual minority | 9 (13.6) |
| Multiple identities | 3 (4.5) |
| Missing | 1 (1.5) |
| Gender identity (N = 64) |  |
| Cisgender male | 21 (31.8) |
| Cisgender female | 19 (28.8) |
| Transgender male | 6 (9.1) |
| Transgender female | 13 (19.7) |
| Gender Queer | 2 (3.0) |
| Multiple identities | 3 (4.5) |
| Missing | 2 (3.0) |
| Military branch (N = 66) |  |
| Army | 37 (56.1) |
| Air Force | 8 (12.1) |
| Navy | 13 (19.7) |
| Marines | 6 (9.1) |
| Coast Guard | 2 (3.0) |
|  | *M (SD)* |
| Age (N = 65) | 47.06 (13.74) |
| Pre-program internalized prejudice (N = 61) | 7.21 (4.73) |
| Post-program internalized prejudice (N = 60) | 6.69 (4.04) |
| Pre-program depressive symptoms (N = 60) | 10.77 (7.03) |
| Post-program depressive symptoms (N = 57) | 9.18 (6.23) |
| Pre-program anxiety symptoms (N = 60) | 8.76 (6.20) |
| Post-program anxiety symptoms (N = 57) | 7.33 (5.20) |
| Pre-program suicide attempt likelihood (N = 66) | 1.54 (1.49) |
| Post-program suicide attempt likelihood (N = 62) | 1.00 (1.25) |
| Problem-solving coping self-efficacy (N = 66) | 37.24 (13.88) |
| Thought stopping coping self-efficacy (N = 66) | 20.46 (10.58) |
| Getting social support coping self-efficacy (N = 66) | 15.24 (8.50) |

Notes. N = Sample size; M = Mean; SD = Standard deviation.

### Procedure

The PRIDE in All Who Served program was funded by the VHA Innovation Ecosystem as an innovation investment program. From October 2017 to September 2019, ten VHA sites participated in data collection efforts for program evaluation and quality improvement purposes. Procedures were reviewed by the Tuscaloosa VA Medical Center Institutional Review Board (Project title: [1316792-1] *Serving All Who Served: Improving Access to Health Care for LGBT Veterans*; IRB Reference #: 00254/19-01) and the VA Central Office program sponsor consistent with federal regulations (e.g., VA Program Guide 1200.21: VHA Operations Activities That May Constitute Research). LGBTQ+ veterans were identified by group facilitators at each site via self-referrals and referrals from other VA providers. A pre/post single group design was used to assess veteran-level changes across the 10-week program. Veteran participants at each site completed a voluntary paper and pencil questionnaire during the first group session (session 1) and last group session (session 10). Informed consent was completed as a verbal discussion in a group setting (during the 1^st^ and 10^th^ sessions of the 10-week group), with written instructions presented on the first page of the paper survey packets. Outcome questionnaires were collected anonymously using a participant-created ID consisting of letters and numbers to connect assessments over time. Questionnaires were distributed and collected by group facilitators from each site who then scanned and returned them to the evaluation team via encrypted email. Participants who only completed the pre- or post-questionnaire were excluded from analyses.

### Measures

#### Demographics

Participants completed a short demographic form which included information such as sexual orientation, gender identity, sex assigned at birth, age, race, ethnicity, and military service branch. Extensive gender and sexual orientation identity label list choices were provided along with the option to write-in responses. In this way, LGBTQ+ veterans were able to express their unique demographic-related identities within the scope of participating in PRIDE in All Who Served (see 2, 3 for more information about the evaluation approach).

#### Coping self-efficacy

The Coping Self-Efficacy Scale (CSES; 35) is a 13-item self-report questionnaire which assesses confidence in engaging in three forms of coping: (a) problem-focused coping (e.g., finding solutions), (b) stopping unpleasant thoughts/emotions (e.g., keeping from feeling sad), and (c) using social support (e.g., making new friends). Items are rated on an 11-point Likert scale, ranging from 0 (*cannot do at all*) to 10 (*certain can do*). Subscale scores are generated for each subcomponent via summation: problem-focused coping (total of 6 items), thought stopping (total of 4 items) and social support (3 items); higher scores indicate greater confidence in coping. Subscale internal consistencies among a prior active-duty military sample ranged from good to excellent (problem-focused coping: *α* = .94; thought stopping: *α* = .89; social support: *α* = .82; (42). Internal consistency in the current study was also high (problem-focused coping: α = .92; thought stopping: α = .94; social support: α = .82).

#### Depressive symptoms

The Patient Health Questionnaire-9 (PHQ-9; 50) is a 9-item self-report questionnaire used to assess DSM-IV depression criteria, such as lack of energy and difficulty sleeping. Items are scored on a 4-point Likert scale ranging from 0 (*not at all*) to 3 (*nearly every day*). Total scores are generated via summation, with higher scores indicating greater presence of depressive symptoms. Scores range from 0 to 27, with the following classifications: 0-4: minimal; 5-9: mild; 10-14: moderate; 15-19: moderately severe; and 20-27: severe. A clinical cut-off score can also be used, with scores greater than 10 indicating clinically elevated risk of major depression. In the current study, the total score was used. The PHQ demonstrates good internal consistency veteran samples (*α* = .86; 54); the internal consistency in the current study was also acceptable for pre-(*α* = 88) and post-program (*α* = 88) scores.

#### Anxiety symptoms

The Generalized Anxiety Disorder-7 (GAD-7; 51) is a 7-item measure of symptoms of generalized anxiety disorder, such as feeling nervous, anxious, or on edge. Items are scored on a 4-point Likert scale ranging from 0 (*not at all*) to 3 (*nearly every day*). Total scores are generated via summation, with higher scores indicating greater presence of anxiety symptoms. Scores range from 0 to 21, with the following classifications: 0-4: minimal; 5-9: mild; 10-14: moderate; 15-21: severe. A clinical cut-off score can also be used, with scores greater than 10 indicating clinically elevated risk of generalized anxiety. In the current study, the total score was used. The GAD-7 demonstrates good internal consistency among veteran samples (α = .89; 55). The internal consistency in the current study was acceptable for pre-(α = .90) and post-program (α = .90) scores.

#### Suicidal attempt risk

The Suicide Behaviors Questionnaire-Revised (SBQ-R; 52) is a 4-item measure of suicidality, including: (a) lifetime suicidal thoughts and behaviors, (b) 12-month suicidal ideation, (c) communication of suicidal intent; and (d) likelihood of future suicidal behavior. Total scores are generated via summation, ranging from 3 to 18, with higher scores indicating higher suicidality. A clinical cut-off score can be generated, with scores greater than or equal to 7 representing clinically significant suicide risk. SBQ-R single items are also frequently used individually in order to assess unique aspects of suicidal behavior. In the current study, the last item concerning suicide attempt likelihood was used. The SBQ-R demonstrates excellent internal consistency among veteran samples (*α* = .94; 56). We did not tabulate internal consistency for the single item used in the current study.

#### Internalized Prejudice

The internalized prejudice subscale of the Lesbian, Gay, and Bisexual Identity Scale (LGBIS; 53) is a 27-item measure quantifying 7 aspects of sexual minority identity. For PRIDE in All Who Served program development, the LGBIS was modified to be inclusive of gender diversity through altered instructions. Specifically, the following wording was added: “For those identifying as heterosexual but as gender diverse, please respond to the items on this page only as you feel comfortable in how they may apply to you” (2, p. 493). Items are rated on a six-point Likert scale ranging from 1 (*disagree strongly*) to 6 (*agree strongly*) (53). The internalized prejudice subscale comprises three items. Internal consistency values were acceptable for pre-(α = .89) and post-program (α = .84) scores.

## Data Analysis

Prior to demographic moderation analyses, race, sexual orientation, and gender identity were recoded due to small cell sizes. We acknowledge that collapsing individuals into larger categories is problematic and potentially invalidating; retaining more categories is preferred (e.g., see 57). The demographics (Table 1) describes the study sample in more detail than was feasible in moderation analyses. Race was reclassified as White (*n* = 31, 47.0%) and racial minority (*n* = 35, 53.0%). In line with sexual identity label literature (e.g., 58, 59), sexual orientation was regrouped as heterosexual/gay/lesbian (HGL; i.e., attraction to one sex; *n* = 34, 51.5%) and bisexual/questioning/and other (BQ+; i.e., combining those to identify in ways other than single-sex attraction; *n* = 31, 47.0%). Two step-gender (i.e., items assessing sex assigned at birth and gender identity) was recoded as cisgender male (*n* = 21, 31.8%), cisgender female (*n* = 19, 28.8%), and transgender/gender diverse (*n* = 24, 36.4%) for the moderation analyses. Repeated measures general linear model (GLM) analyses (60, 61) were used to examine program moderation analyses. Repeated-measures GLM was selected because the approach: (a) allows for simultaneous inclusion of categorical and continuous variables; (b) enables tests of pre-post program main effects, as well as baseline by pre-post program outcome interaction terms; and (c) provides a flexible set of multivariate and univariate test statistics for interpretation. For all models, the pre-post program outcome was either a mental health symptom (e.g., depressive symptoms) or internalized prejudice. For all models, the moderator was either a coping self-efficacy subscale or demographic variable (e.g., age). For example, for an age by depressive symptoms analysis, the independent variable is the pre-post change in symptoms of depressive symptoms and the moderator is age. We used multivariate (i.e., Wilk’s λ) omnibus tests statistics because doing so reduces likelihood of Type I error. We utilized guidelines for magnitude of effect size interpretation for partial-eta squared values provided by Cohen (62): small = .01, medium = .06, large = .14. Significant interactions were graphed following procedures recommended by Bauer and Curran (63), including representing low and high levels of continuous moderators at -/+ one standard deviation around the mean.

## Results

### Coping Self-Efficacy as a Moderator of PRIDE in All Who Served Program Outcomes

Table 2 contains test statistics for all CSE subscale moderation analyses. Each CSE subscale was examined across four separate repeated measures GLM analyses, one for each outcome of interest (i.e., depressive symptoms, anxiety symptoms, suicide attempt likelihood, and internalized prejudice). Reviewing Table 2, problem-solving CSE moderated pre-post program reductions in depressive symptoms, anxiety symptoms, and internalized prejudice (all medium effects). Figures 1a through 1c depict the interactions. As predicted, LGBTQ+ veterans lowest in problem-solving CSE reported greater reductions in depressive symptoms and internalized prejudice after attending the PRIDE in All Who Served group. Additionally, LGBTQ+ veterans with moderate problem-solving CSE also reported reductions in depressive and anxiety symptoms.

**Table 2.** Coping self-efficacy subscales as a moderator of treatment effects using repeated measures general linear modeling.

| Predictor | Depressive Symptoms |  |  |  | Anxiety Symptoms |  |  |  | Suicide Attempt Likelihood |  |  |  | Internalized Prejudice |  |  |  |
| --- | --- | --- | --- | --- | --- | --- | --- | --- | --- | --- | --- | --- | --- | --- | --- | --- |
| | Wilks' $\lambda$ | F (df) | p-value | $\eta_p^2$ | Wilks' $\lambda$ | F (df) | p-value | $\eta_p^2$ | Wilks' $\lambda$ | F (df) | p-value | $\eta_p^2$ | Wilks' $\lambda$ | F (df) | p-value | $\eta_p^2$ |
| Time | 0.84 | 10.04 (1, 55) | .003 | .15*** | 0.86 | 9.04 (1, 55) | .004 | .14*** | 0.88 | 7.98 (1, 60) | .006 | .12** | 0.89 | 7.36 (1, 59) | .009 | .11** |
| CSE Problem-solving by time | **0.91** | **5.51 (1, 55)** | **.023** | **.09\*\*** | **0.92** | **4.69 (1, 55)** | **.035** | **.08\*\*** | 0.96 | 2.69 (1, 60) | .106 | .04* | **0.93** | **4.39 (1, 59)** | **.040** | **.07\*\*** |
| Time | 0.90 | 6.03 (1, 55) | .017 | .10** | 0.87 | 7.87 (1, 55) | .007 | .12** | 0.83 | 12.25 (1, 60) | .001 | .17*** | 0.86 | 9.76 (1, 59) | .003 | .14*** |
| CSE Thought Stopping by time | 0.97 | 1.82 (1, 55) | .183 | .03* | 0.95 | 2.91 (1, 55) | .094 | .05* | 0.94 | 3.90 (1, 60) | .053 | .06** | **0.91** | **5.64 (1, 59)** | **.021** | **.09\*\*** |
| Time | 0.82 | 12.24 (1, 55) | .001 | .18*** | 0.85 | 9.74 (1, 55) | .003 | .15*** | 0.82 | 13.08 (1, 60) | .001 | .18*** | 0.86 | 9.60 (1, 59) | .003 | .14*** |
| CSE Social Support by time | **0.90** | **5.84 (1, 55)** | **.019** | **.10\*\*** | 0.94 | 3.94 (1, 55) | .052 | .07** | **0.94** | **4.03 (1, 60)** | **.049** | **.06\*\*** | **0.92** | **5.36 (1, 59)** | **.024** | **.08\*\*** |

Notes: \* = Small effect, \*\* = Medium effect, \*\*\* = Large effect per Cohen (1998) guidelines. **Bold font** denotes **significant moderation effect**, significant main effects of time on each outcome are not bolded. CSE = Coping self-efficacy; Time = Change in pre-to-post program scores;; $\eta_p^2$ = Partial eta-squared; A total of our separate models were run for each CSE subscale, each having a time main effect and one interaction term.

**Figures 1a to 1c.**
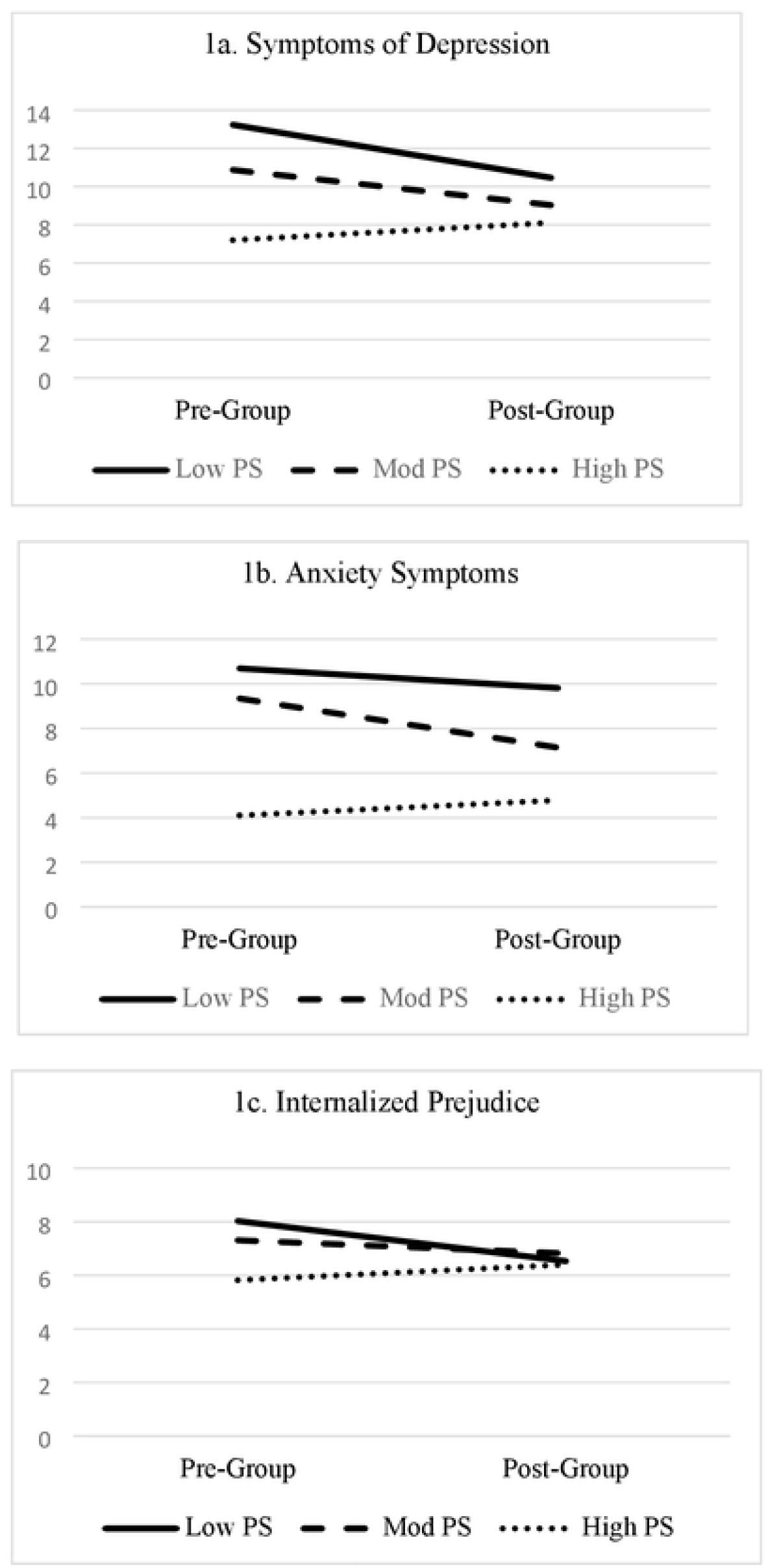
Problem-solving coping-self efficacy as a significant moderator of treatment effect on symptoms of depression, anxiety symptoms, and internalized prejudice. Notes. Depression= Patient Health Questionnaire (PHQ-9); Anxiety= Generalized Anxiety Disorder (GAD-7); IP= internalized prejudice subscale of the Lesbian, Gay, and Bisexual Identity Scale (LGBIS); PS= Problem-solving subscale of the Coping Self-Efficacy scale. Test statistics in Table 2.

Referencing Table 2, thought stopping CSE moderated pre-post reductions in internalized prejudice (medium effect). Figure 2 depicts the thought stopping CSE by internalized prejudice interaction. LGBTQ+ veterans with moderate thought stopping CSE reported reductions in internalized prejudice. An unanticipated pattern emerged in that LGBTQ+ veterans with high thought stopping CSE reported increased internalized prejudice.

**Figure 2.**
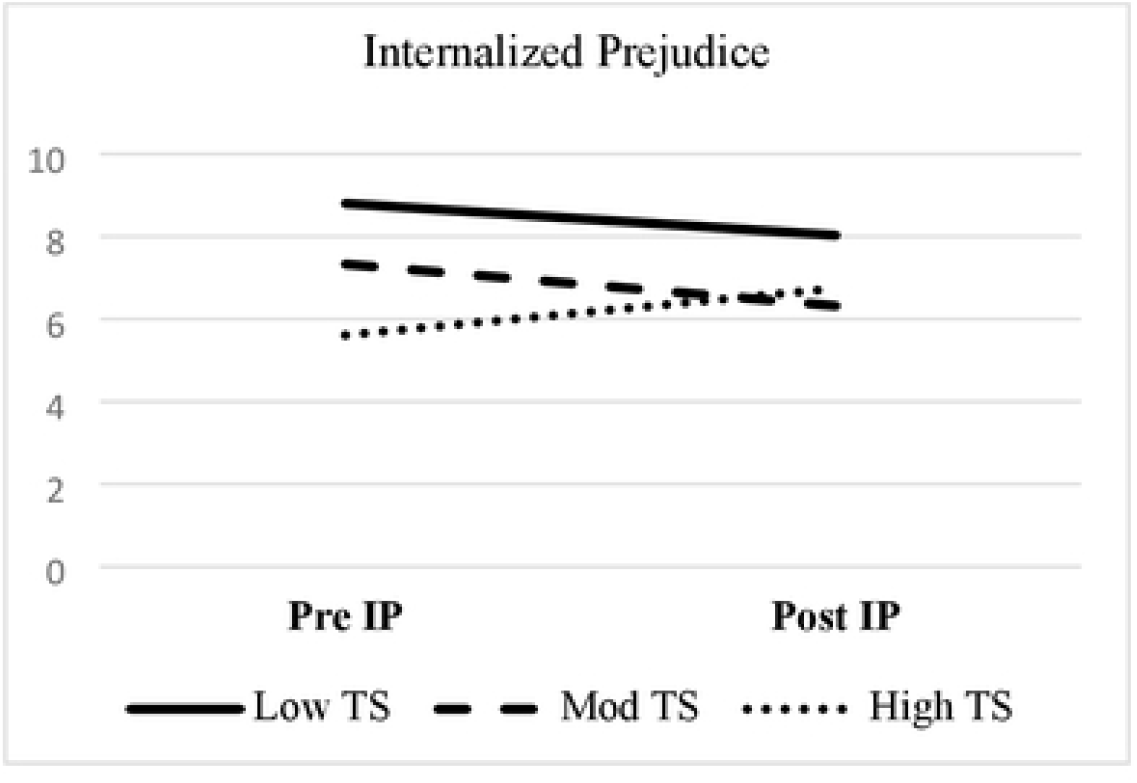
Thought stopping coping-self efficacy as a significant moderator of internalized prejudice. Notes. TS = Thought stoppingsubscale of the Coping Self-Efficacy scale;IP= internalized prejudice subscale of the Lesbian, Gay, and Bisexual Identity Scale (LGBIS). Test statistics presented in Table 2.

Also in Table 2, social support CSE moderated pre-post reductions in depressive symptoms, suicide attempt likelihood, and internalized prejudice (all medium effects). Figures 3a to 3c depicts the interactions. In line with expectations, LGBTQ+ veterans low in social support CSE reported greater reductions in depressive symptoms, suicidal attempt likelihood, and internalized prejudice after attending the group. Also, LGBTQ+ veterans moderate in social support CSE reported reductions in depressive symptoms, suicidal attempt likelihood, and internalized prejudice. An unanticipated pattern emerged in that LGBTQ+ veterans high in social support CSE reported increased internalized prejudice.

**Figures 3a to 3c.**
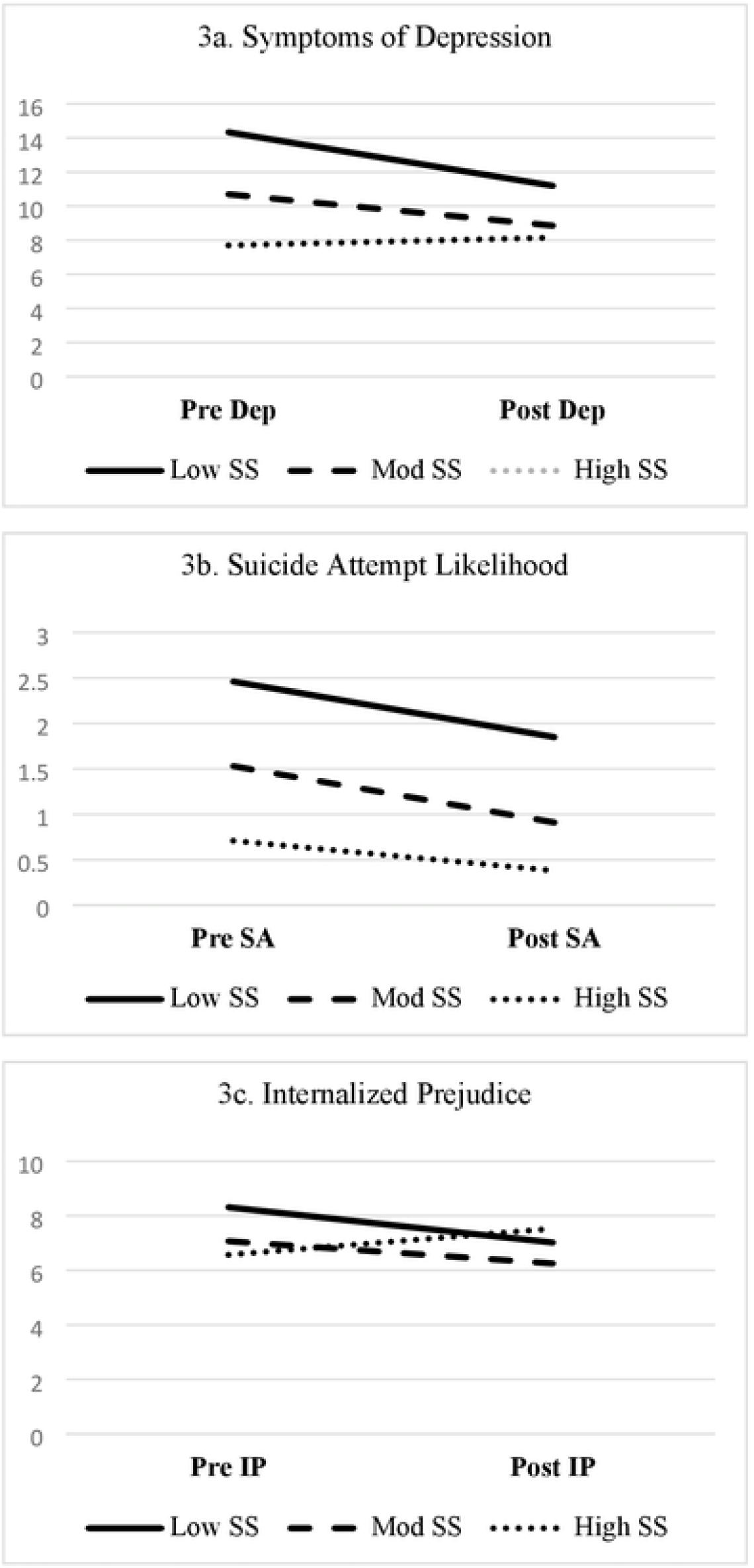
Social support copingse f efficacy as a significant moderator of symptoms of depression, suicide attempt likelihood, and internalized prejudice. Notes. Dep= Symptoms of depression; SA= suicide attempt likelihood; IP= internalized prejudice subscale of the Lesbian, Gay, and Bisexual Identity Scale (LGBIS); SS =Soc al support subscale of the Coping Sel fEfficacy scale. Test stat stics in Table 2.

### Demographics as Moderators of PRIDE in All Who Served Program Outcomes

Table 3 contains demographic moderation analysis test statistics. The only observed significant moderation effect was sexual orientation on pre-post reductions in internalized prejudice (medium effect). Figure 4 depicts this interaction. HGL veterans reported no change in internalized prejudice, whereas BQ+ veterans reported reductions in internalized prejudice.

**Table 3.** Moderation analyses using repeated measures general linear modeling for demographic characteristics.

| Predictor | Depressive Symptoms |  |  |  | Anxiety Symptoms |  |  |  | Suicide Attempt Likelihood |  |  |  | Internalized Prejudice |  |  |  |
| --- | --- | --- | --- | --- | --- | --- | --- | --- | --- | --- | --- | --- | --- | --- | --- | --- |
| | Wilks' $\lambda$ | F (df) | p-value | $\eta_p^2$ | Wilks' $\lambda$ | F (df) | p-value | $\eta_p^2$ | Wilks' $\lambda$ | F (df) | p-value | $\eta_p^2$ | Wilks' $\lambda$ | F (df) | p-value | $\eta_p^2$ |
| Time | 0.94 | 3.32 (1, 54) | .074 | .06** | 0.97 | 1.88 (1, 54) | .176 | .03* | 0.93 | 4.46 (1, 59) | .039 | .07** | 0.98 | 1.25 (1, 58) | .268 | .02* |
| Age by time | 0.98 | 1.29 (1, 54) | .260 | .02* | 0.99 | 0.46 (1, 54) | .500 | .01* | 0.98 | 1.29 (1, 59) | .261 | .02* | 0.99 | 0.31 (1, 58) | .579 | .005* |
| Time | 0.88 | 7.52 (1, 55) | .008 | .12** | 0.88 | 7.75 (1, 55) | .007 | .12** | 0.82 | 13.14 (1, 60) | .001 | .18*** | 0.93 | 4.38 (1, 59) | .041 | .07** |
| Race by time | 0.94 | 3.34 (1, 55) | .073 | .06** | 0.94 | 3.35 (1, 55) | .073 | .06** | 0.99 | 0.36 (1, 60) | .553 | .01* | 1.00 | 0.08 (1, 59) | .778 | .001* |
| Time | 0.87 | 8.05 (1, 54) | .006 | .13** | 0.88 | 7.07 (1, 54) | .010 | .12** | 0.82 | 12.57 (1, 59) | .001 | .18*** | 0.91 | 5.78 (1, 58) | .019 | .09** |
| Sexual Orientation by time | 0.96 | 1.95 (1, 54) | .169 | .03* | 1.00 | 0.01 (1, 54) | .936 | .00* | 0.99 | 0.43 (1, 59) | .513 | .01* | **0.91** | **5.36 (1, 58)** | **.024** | **.08\*\*** |
| Time | 0.89 | 6.08 (1, 52) | .017 | .10** | 0.88 | 6.93 (1, 52) | .011 | .12** | 0.83 | 11.93 (1, 57) | .001 | .17*** | 0.95 | 3.10 (1, 56) | .084 | .05* |
| Gender Identity by time | 0.97 | 0.68 (1, 52) | .511 | .03* | 1.00 | 0.06 (1, 52) | .940 | .00* | 0.98 | 0.53 (1, 57) | .593 | .02* | 0.98 | 0.57 (1, 56) | .567 | .02* |

Notes: \* = Small effect, \*\* = Medium effect, \*\*\* = Large effect per Cohen (1998) guidelines. **Bold font** denotes **significant moderation effect**.
Race = White or racial minority; Sexual orientation = Heterosexual/gay/lesbian or bisexual/queer/questioning/other; Gender = Cisgender male, cisgender female, or transgender/gender diverse; Time = Change in pre-to-post program scores at 10 weeks; $\eta_p^2$ = Partial eta-squared. A separate model was run for each CSE subscale, such that each has a main effect of time (pre-post group change) and one interaction term.

**Figure 4.**
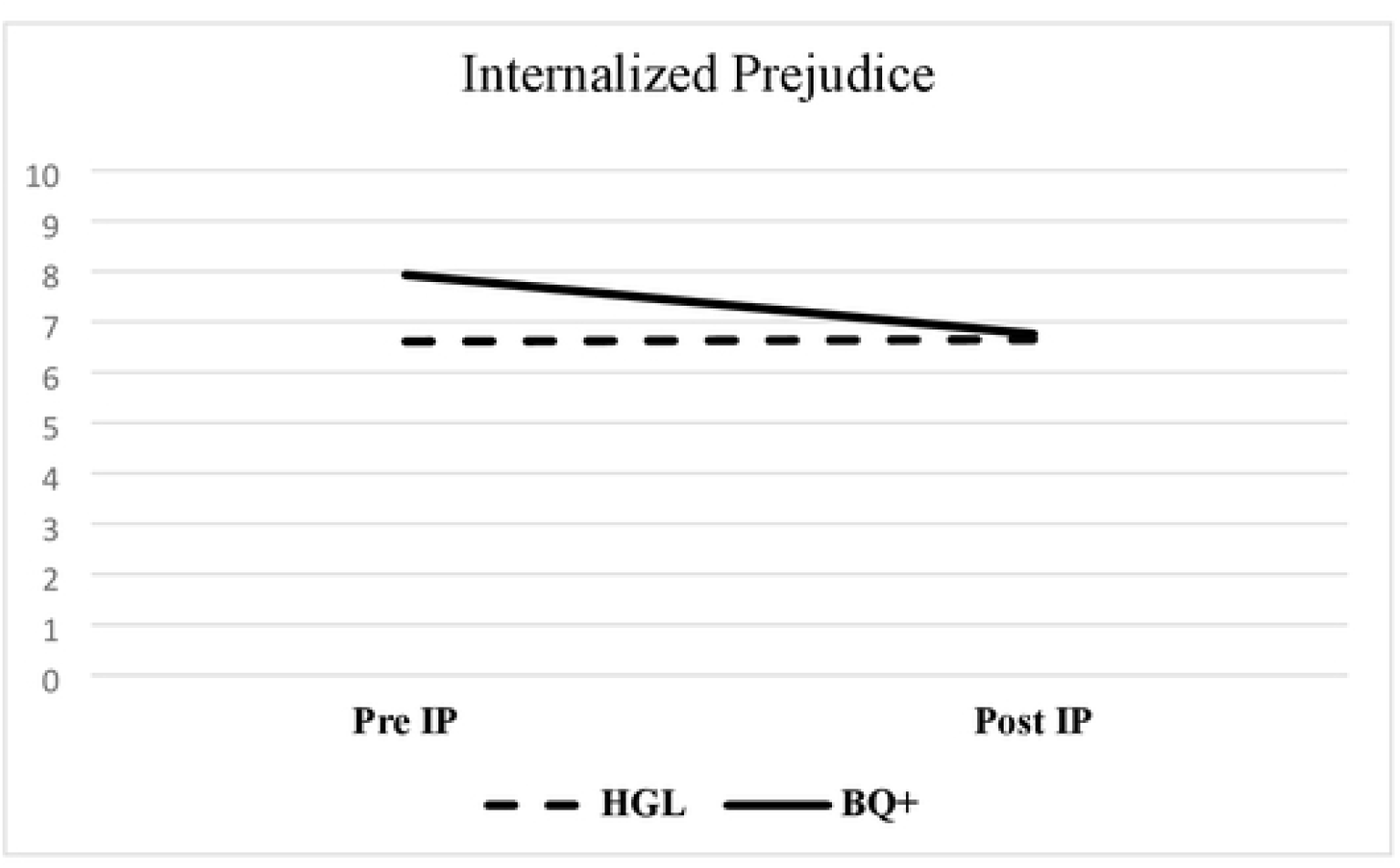
Sexual orientation as a significant moderator of internalized prejudice. Notes.IP= internalized prejudice subscale of the Lesbian, Gay, and Bisexual Identity Scale (LGBIS); HGL = Heterosexual/gay/lesbian; BQ+ = Bisexual/queer/questioning/somethingelse. Test statistics in Table 3.

## Discussion

As predicted, LGBTQ+ veterans possessing low, and at times moderate, levels of CSE experienced the largest improvements in mental health and internalized stigma outcomes during the PRIDE in All Who Served program. LGBTQ+ veterans with low or moderate problem-solving CSE reported reductions in symptoms of depression, anxiety, and internalized prejudice. Moreover, LGBTQ+ veterans with low or moderate degrees of social support CSE saw reductions in symptoms of depression, internalized prejudice, and suicide attempt likelihood. The moderating role of coping-related beliefs is consistent with the Minority Stress Model (10, 11) premise that coping and connectedness can buffer identity- and discrimination-related impacts on mental health and well-being. Our findings are also consistent with cross-sectional research showing (a) stress-buffering effects of coping (e.g., using social support) among LGBTQ+ people (e.g., 25, 26) and (b) a pattern of negative associations between CSE beliefs and severity of symptoms of mental illness among LGBTQ+ people (e.g., 40, 41).

From a CSE perspective, our findings are not consistent with preliminary tests of CSE and suicidality among active-duty military personnel. In particular, Cunningham and colleagues (42) identified CSE thought stopping beliefs as a primary factor implicated in suicide risk. Bowling and colleagues (64) observed that (a) transgender and non-binary adults possessed lower CSE thought stopping compared to other genders and (b) all sexual minority adults in their sample demonstrated lower CSE thought stopping compared to heterosexual counterparts. Prior research therefore illustrates the potential primacy of CSE thought stopping in understanding suicide among military personnel, and between-groups variation by LGBTQ+ identity. On the contrary, we observed primacy of CSE problem-solving and social support for LGBTQ+ veterans, suggesting that varying aspects of CSE may be more beneficial or relevant to cisgender/heterosexual versus LGBTQ+ military-affiliated personnel.

CSE problem-solving and social support belief moderation patterns raise the question of possible mechanisms by which these findings can be contextualized. The PRIDE in All Who Served program (2) is one of a broader set of affirmative group-based interventions for LGBTQ+ persons emerging in the literature (e.g., 65, 66). Recent qualitative inquiries into the benefits of affirmative cognitive behavior therapy (CBT)-based group interventions in particular (e.g., 67, 68) may shed light on why low-to-moderate CSE beliefs are associated with PRIDE program gains. Sexual minority persons participating in affirmative CBT group interventions have suggested that engagement with CBT tools and concepts (68), as well as the decreased sense of loneliness from interaction with other sexual minority men of color (67), may explain improved mental health. Applied to our findings, LGBTQ+ veterans with lower levels of CSE social support and problem-solving beliefs (a CBT coping skill) may have experienced gains in these areas, thereby affecting positive well-being outcomes. Future research could be conducted to engage group participants in mixed-method inquiry around plausible mechanisms of action to refine our understanding for future PRIDE implementation and other health promotion interventions.

Our CSE moderation analyses showed some indication that those high in CSE thought stopping and social support beliefs may experience slight increases in internalized prejudice. This pattern is contrary to what we expected. While this pattern should be probed for further understanding, we also avoid over emphasis of the trend, particularly given the low levels of internalized prejudice reported at both pre- and post-group timepoints. At least two plausible explanations exist. First, to the extent those high in CSE beliefs at intervention baseline are also quite low in internalized prejudice, the pattern over the course of the PRIDE program may reflect some regression to the mean. Alternatively, characteristics of the group may have affected levels of internalized prejudice. Lloyd and colleagues (68) observed that some sexual minority adults taking part in an adapted CBT group therapy program found generational divides between participant groups to harm group cohesion.

Overall, demographics did not moderate program outcomes, suggesting that an inclusive health promotion group – as originally developed – is likely a good match for the heterogenous individuals typically enrolled. One notable exception was observed for veterans who reported their sexual orientations as bisexual, queer, questioning or something else (BQ+, e.g., pansexual, see Table 1). BQ+ veterans in this sample reported greater reduction in internalized prejudice following the group than their single-gender attracted peers (i.e., heterosexual, gay, or lesbian veterans). Though based on a relatively small sample and conducted within a clinical setting rather than a rigorous clinical trial, the data presented here offer preliminary evidence of an intervention with a promising impact for BQ+ individuals consistent with recent calls to action for the field (69). The unique experiences of discrimination, dismissal, and invisibility of bisexual people including lack of in-group community support has been reported in both qualitative and larger quantitative/epidemiological studies (e.g., 70, 71). For example, individuals who are attracted to more than one gender may experience discrimination from both heterosexual individuals as well as from within the LGBTQ+ community. Our findings are of course contextualized by the limited number of individuals in the current sample, some attrition from pre- to post-assessment, and the limited time frame for follow-up (e.g., outcomes at 10-weeks). Expansion to include more individuals who identity as racial and ethnic minorities, a larger sample that would permit more in-depth analyses of gender (57), and exploration of the longevity of these findings is necessary in future work.

## Conclusions

As more healthcare services are developed to address health inequities for historically marginalized people, papers like this one may advance understanding of individual characteristics (e.g., coping self-efficacy, BQ+ identity) not widely considered in clinical care. This paper provides an initial examination of potential moderators of treatment outcomes for LGBTQ+ veterans attending a health promotion group at VA Medical Centers that may inform future tailoring of programs that build resilience, nurture a sense of community, and ultimately provide tangible, upstream suicide prevention strategies. Although the absence of moderation in a small sample is not conclusive, it also provides preliminary program evaluation data to clinicians and policy makers implementing programs for LGBTQ+ veterans. For example, settings are tempted to separate LGBQ and transgender/non-binary veterans into separate groups. However, this group segregation could bring risks (e.g., a veterans who had not historically identified as gender diverse may begin to identify with information about gender outside of a gender binary). Anecdotally, clinicians delivering the group also report rich discussions between younger and older veterans, those with different racial identities (e.g., Black and White individuals) and along other intersections of identity (e.g., religious affiliation, geographic rurality) in the same group. Overall, this study provides an understanding of how LGBTQ+ Veterans with more coping deficits can especially benefit from the PRIDE in All Who Served program.

## Data Availability

The data underlying the results presented in the paper are available from the first author pending acceptable data use agreements.

## Acknowledgments/ Funding

This work was supported by a series of investments from the VHA Innovation Ecosystem, including Seed and Spread Awards (Lange & Hilgeman, Co-Leads, 2017-2019) and Diffusion of Excellence Support (Lange & Hilgeman, Co-Leads 2020-2021, and Sperry & Hilgeman, Co-Leads 2021-2023). Dr. Wilson is supported by IK2HX002398. We deeply appreciate the commitment and ongoing partnership with Blaine Fitzgerald, Diffusion Specialist; PRIDE in All Who Served Group facilitators at early adopting sites; and Operations Partners including the VHA Innovation Network, Diffusion of Excellence, the Lesbian, Gay, Bisexual, Transgender, Queer/Questioning (LGBTQ+) Health Program Office, and the Office of Health Equity.

## Footnotes

1 Note, we are using language reported in original papers to describe research samples (i.e., “men” and “women” rather than “cisgender men” and “cisgender women”) where gender identity and experience are not recorded.

## References

(1) Kazdin AE. Progression of therapy research and clinical application of treatment require better understanding of the change process. Clinical Psychology: Science and Practice. 2001;8:143–51.

(2) Lange TM, Hilgeman MM, Portz KJ, Intoccia VA, Cramer RJ. Pride in all Who Served: Development, Feasibility, and Initial Efficacy of a Health Education Group For LGBT Veterans. J Trauma Dissociation. 2020;21(4):484–504.

(3) Hilgeman MM, Lange TM, Bishop T, Cramer RJ. Spreading pride in all who served: A health education program to improve access and mental health outcomes for sexual and gender minority veterans. Psychol Serv. 2022.

(4) McGirr J, Jones K, Moy E. Chartbook on the Health of Lesbian, Gay, & Bisexual Veterans. In: Office of Health Equity VAHA, U.S. Department of Veterans Affairs, editor. 2021.

(5) Office USGA. VA Health Care: Better Data Needed to Assess the Health Outcomes of Lesbian, Gay, Bisexual, and Transgender Veterans (GAO-21-69). 2020.

(6) Meadows SO, Engel CC, Collins RL, Beckman RL, Cefalu M, Hawes-Dawson J, et al. 2015 Department of Defense Health Related Behaviors Survey (HRBS). Rand Health Q. 2018;8(2):5.

(7) Livingston NA, Berke DS, Ruben MA, Matza AR, Shipherd JC. Experiences of trauma, discrimination, microaggressions, and minority stress among trauma-exposed LGBT veterans: Unexpected findings and unresolved service gaps. Psychol Trauma. 2019;11(7):695–703.

(8) Cochran BN, Balsam K, Flentje A, Malte CA, Simpson T. Mental health characteristics of sexual minority veterans. J Homosex. 2013;60(2-3):419–35.

(9) Mattocks KM, Kauth MR, Sandfort T, Matza AR, Sullivan JC, Shipherd JC. Understanding Health-Care Needs of Sexual and Gender Minority Veterans: How Targeted Research and Policy Can Improve Health. LGBT Health. 2014;1(1):50–7.

(10) Meyer IH. Prejudice, social stress, and mental health in lesbian, gay, and bisexual populations: conceptual issues and research evidence. Psychol Bull. 2003;129(5):674–97.

(11) Testa RJ, Habarth J, Peta J, Balsam K, Bockting W. Development of the Gender Minority Stress and Resilience Measure. Psychology of Sexual Orientation and Gender Diversity. 2015;2(1):65–77.

(12) Blosnich JR, Marsiglio MC, Gao S, Gordon AJ, Shipherd JC, Kauth M, et al. Mental Health of Transgender Veterans in US States With and Without Discrimination and Hate Crime Legal Protection. Am J Public Health. 2016;106(3):534–40.

(13) Tucker RP, Testa RJ, Reger MA, Simpson TL, Shipherd JC, Lehavot K. Current and Military-Specific Gender Minority Stress Factors and Their Relationship with Suicide Ideation in Transgender Veterans. Suicide Life Threat Behav. 2019;49(1):155–66.

(14) Don’t Ask, Don’t Tell, 10 U.S.C. § 654. 1993.

(15) Don’t Ask, Don’t Tell Repeal Act of 2010, H.R.2965. 2009.

(16) Defense USDo. Secretary of Defense Ash Carter Announces Policy for Transgender Service Members. 2016.

(17) House TW. Executive Order on Enabling All Qualified Americans to Serve Their Country in Uniform. 2021.

(18) McNamara KA, Lucas CL, Goldbach JT, Castro CA, Holloway IW. “Even If the Policy Changes, the Culture Remains the Same”: a mixed methods analysis of LGBT service members’ outness patterns. Armed Forces & Society. 2021;47(3):505–29.

(19) Feinstein BA, Xavier Hall CD, Dyar C, Davila J. Motivations for sexual identity concealment and their associations with mental health among bisexual, pansexual, queer, and fluid (bi+) individuals. J Bisex. 2020;20(3):324–41.

(20) Pachankis JE. The psychological implications of concealing a stigma: a cognitive-affective-behavioral model. Psychol Bull. 2007;133(2):328–45.

(21) Excellence Do. PRIDE In All Who Served: Diffusion Marketplace; 2022 [Available from: https://marketplace.va.gov/innovations/pride-in-all-who-served-reducing-healthcare-disparities-for-lgbt-veterans.

(22) Bucher MA, Suzuki T, Samuel DB. A meta-analytic review of personality traits and their associations with mental health treatment outcomes. Clinical Psychology Review. 2019;70:51–63.

(23) Hilgeman MM, Allen RS, DeCoster J, Burgio LD. Positive aspects of caregiving as a moderator of treatment outcome over 12 months. Psychol Aging. 2007;22(2):361–71.

(24) Rosenblum A, Foote J, Cleland C, Magura S, Mahmood D, Kosanke N. Moderators of Effects of Motivational Enhancements to Cognitive Behavioral Therapy. The American Journal of Drug and Alcohol Abuse. 2005;31(1):35–58.

(25) McConnell EA, Janulis P, Phillips G, 2nd, Truong R, Birkett M. Multiple Minority Stress and LGBT Community Resilience among Sexual Minority Men. Psychol Sex Orientat Gend Divers. 2018;5(1):1–12.

(26) Rogers AH, Seager I, Haines N, Hahn H, Aldao A, Ahn WY. The Indirect Effect of Emotion Regulation on Minority Stress and Problematic Substance Use in Lesbian, Gay, and Bisexual Individuals. Front Psychol. 2017;8:1881.

(27) Toomey RB, Ryan C, Diaz RM, Russell ST. Coping With Sexual Orientation-Related Minority Stress. J Homosex. 2018;65(4):484–500.

(28) Aranda F, Matthews AK, Hughes TL, Muramatsu N, Wilsnack SC, Johnson TP, et al. Coming out in color: racial/ethnic differences in the relationship between level of sexual identity disclosure and depression among lesbians. Cultur Divers Ethnic Minor Psychol. 2015;21(2):247–57.

(29) Balsam KF, Molina Y, Blayney JA, Dillworth T, Zimmerman L, Kaysen D. Racial/ethnic differences in identity and mental health outcomes among young sexual minority women. Cultur Divers Ethnic Minor Psychol. 2015;21(3):380–90.

(30) Griner D, Smith TB. Culturally adapted mental health intervention: A meta-analytic review. Psychotherapy (Chic). 2006;43(4):531–48.

(31) Rojas-García A, Ruiz-Perez I, Rodríguez-Barranco M, Gonçalves Bradley DC, Pastor-Moreno G, Ricci-Cabello I. Healthcare interventions for depression in low socioeconomic status populations: A systematic review and meta-analysis. Clin Psychol Rev. 2015;38:65–78.

(32) Smith TB, Rodríguez MD, Bernal G. Culture. Journal of Clinical Psychology. 2011;67(2):166–75.

(33) Beck JG, Clapp JD, Unger W, Wattenberg M, Sloan DM. Moderators of PTSD symptom change in group cognitive behavioral therapy and group present centered therapy. J Anxiety Disord. 2021;80:102386.

(34) Cason D, Grubaugh A, Resick P. Gender and PTSD treatment: Efficacy and effectiveness. In: Kimerling R, Ouimette P, Wolfe J, editors. Gender and PTSD: The Guilford Press; 2002. p. 305–34.

(35) Chesney MA, Neilands TB, Chambers DB, Taylor JM, Folkman S. A validity and reliability study of the coping self-efficacy scale. Br J Health Psychol. 2006;11(Pt 3):421–37.

(36) Burton CL, Wang K, Pachankis JE. Psychotherapy for the Spectrum of Sexual Minority Stress: Application and Technique of the ESTEEM Treatment Model. Cogn Behav Pract. 2019;26(2):285–99.

(37) Flentje A. AWARENESS: Development of a cognitive-behavioral intervention to address intersectional minority stress for sexual minority men living with HIV who use substances. Psychotherapy (Chic). 2020;57(1):35–49.

(38) Matsuno E, Israel T. Psychological Interventions Promoting Resilience Among Transgender Individuals: Transgender Resilience Intervention Model (TRIM). The Counseling Psychologist. 2018;46(5):632–55.

(39) Goldin PR, Morrison A, Jazaieri H, Brozovich F, Heimberg R, Gross JJ. Group CBT versus MBSR for social anxiety disorder: A randomized controlled trial. J Consult Clin Psychol. 2016;84(5):427–37.

(40) Cramer RJ, Burks AC, Plöderl M, Durgampudi P. Minority stress model components and affective well-being in a sample of sexual orientation minority adults living with HIV/AIDS. AIDS Care. 2017;29(12):1517–23.

(41) Denton FN, Rostosky SS, Danner F. Stigma-related stressors, coping self-efficacy, and physical health in lesbian, gay, and bisexual individuals. J Couns Psychol. 2014;61(3):383–91.

(42) Cunningham CA, Cramer RJ, Cacace S, Franks M, Desmarais SL. The Coping Self-Efficacy Scale: Psychometric properties in an outpatient sample of active duty military personnel. Military Psychology. 2020;32(3):261–72.

(43) Bartone PT. Resilience Under Military Operational Stress: Can Leaders Influence Hardiness? Military Psychology. 2006;18(sup1):S131–S48.

(44) Ramirez MH, Rogers SJ, Johnson HL, Banks J, Seay WP, Tinsley BL, et al. If we ask, what they might tell: clinical assessment lessons from LGBT military personnel post-DADT. J Homosex. 2013;60(2-3):401–18.

(45) Cox DW, Bakker AM, Naifeh JA. Emotion Dysregulation and Social Support in PTSD and Depression: A Study of Trauma-Exposed Veterans. J Trauma Stress. 2017;30(5):545–9.

(46) Han SC, Castro F, Lee LO, Charney ME, Marx BP, Brailey K, et al. Military unit support, postdeployment social support, and PTSD symptoms among active duty and National Guard soldiers deployed to Iraq. J Anxiety Disord. 2014;28(5):446–53.

(47) Tuncay T, Musabak I. Problem-Focused Coping Strategies Predict Posttraumatic Growth in Veterans With Lower-Limb Amputations. Journal of Social Service Research. 2015;41(4):466–83.

(48) Cramer RJ, Kaniuka AR, Lange TM, Brooks BD, Feinstein BA, Hilgeman MM. A pilot evaluation of sexual and gender minority identity measures in a treatment-engaged military veteran sample. Am J Orthopsychiatry. 2022;92(4):442–51.

(49) Kilbourne AM, Evans E, Atkins D. Learning health systems: Driving real-world impact in mental health and substance use disorder research. FASEB Bioadv. 2021;3(8):626–38.

(50) Kroenke K, Spitzer RL, Williams JB. The PHQ-9: validity of a brief depression severity measure. J Gen Intern Med. 2001;16(9):606–13.

(51) Spitzer RL, Kroenke K, Williams JB, Löwe B. A brief measure for assessing generalized anxiety disorder: the GAD-7. Arch Intern Med. 2006;166(10):1092–7.

(52) Osman A, Bagge CL, Gutierrez PM, Konick LC, Kopper BA, Barrios FX. The Suicidal Behaviors Questionnaire-Revised (SBQ-R): validation with clinical and nonclinical samples. Assessment. 2001;8(4):443–54.

(53) Mohr JJ, Kendra MS. Revision and extension of a multidimensional measure of sexual minority identity: the Lesbian, Gay, and Bisexual Identity Scale. J Couns Psychol. 2011;58(2):234–45.

(54) Corson K, Gerrity MS, Dobscha SK. Screening for depression and suicidality in a VA primary care setting: 2 items are better than 1 item. Am J Manag Care. 2004;10(11 Pt 2):839–45.

(55) Ben Barnes J, Presseau C, Jordan AH, Kline NK, Young-McCaughan S, Keane TM, et al. Common Data Elements in the Assessment of Military-Related PTSD Research Applied in the Consortium to Alleviate PTSD. Mil Med. 2019;184(5-6):e218–e26.

(56) Franks M, Cramer RJ, Cunningham CA, Kaniuka AR, Bryan CJ. Psychometric assessment of two suicide screeners when used under routine conditions in military outpatient treatment programs. Psychol Serv. 2021;18(3):433–9.

(57) Bauer GR, Braimoh J, Scheim AI, Dharma C. Transgender-inclusive measures of sex/gender for population surveys: Mixed-methods evaluation and recommendations. PLoS One. 2017;12(5):e0178043.

(58) Galupo MP, Mitchell RC, Davis KS. Sexual minority self-identification: Multiple identities and complexity. Psychology of sexual orientation and gender diversity. 2015;2(4):355.

(59) Galupo MP, Davis KS, Grynkiewicz AL, Mitchell RC. Conceptualization of Sexual Orientation Identity Among Sexual Minorities: Patterns Across Sexual and Gender Identity. Journal of Bisexuality. 2014;14(3-4):433–56.

(60) Cohen J, Cohen P, West SG, Aiken LS. Applied Multiple Regression/Correlation Analysis for the Behavioral Sciences. 3rd ed: Lawrence Erlbaum Associates Publishers; 2003.

(61) Ho R. Handbook of Univariate and Multivariate Data Analysis and Interpretation with SPSS. 1st ed: Chapman and Hall/CRC; 2006.

(62) Cohen J. Statistical Power Analysis Jbr the Behavioral. Sciences Hillsdale (NJ): Lawrence Erlbaum Associates. 1988:18–74.

(63) Bauer DJ, Curran PJ. Probing Interactions in Fixed and Multilevel Regression: Inferential and Graphical Techniques. Multivariate Behav Res. 2005;40(3):373–400.

(64) Bowling J, Montanaro E, Cramer RJ, Mennicke A, Wilsey CN, Kaniuka AR, et al. Gender, sexual orientation, and mental health in the kink community: an application of coping self-efficacy theory. Psychol Health. 2021:1–16.

(65) Millar BM, Wang K, Pachankis JE. The moderating role of internalized homonegativity on the efficacy of LGB-affirmative psychotherapy: Results from a randomized controlled trial with young adult gay and bisexual men. Journal of Consulting and Clinical Psychology. 2016;84(7):565.

(66) Pachankis JE, Soulliard ZA, Morris F, Seager van Dyk I. A Model for Adapting Evidence-Based Interventions to Be LGBQ-Affirmative: Putting Minority Stress Principles and Case Conceptualization Into Clinical Research and Practice. Cognitive and Behavioral Practice. 2022.

(67) Jackson SD, Wagner KR, Yepes M, Harvey TD, Higginbottom J, Pachankis JE. A pilot test of a treatment to address intersectional stigma, mental health, and HIV risk among gay and bisexual men of color. Psychotherapy (Chic). 2022;59(1):96–112.

(68) Lloyd CEM, Rimes KA, Hambrook DG. LGBQ adults’ experiences of a CBT wellbeing group for anxiety and depression in an Improving Access to Psychological Therapies Service: a qualitative service evaluation. the Cognitive Behaviour Therapist. 2021;13:e58.

(69) Taylor J. Bisexual Mental Health: A Call to Action. Issues Ment Health Nurs. 2018;39(1):83–92.

(70) Ghabrial MA, Ross LE. Representation and erasure of bisexual people of color: A content analysis of quantitative bisexual mental health research. Psychology of Sexual Orientation and Gender Diversity. 2018;5:132–42.

(71) Ross LE, Dobinson C, Eady A. Perceived determinants of mental health for bisexual people: a qualitative examination. Am J Public Health. 2010;100(3):496–502.

